# A Note on COVID-19 Diagnosis Number Prediction Model in China

**DOI:** 10.1101/2020.02.19.20025262

**Authors:** Yi Li, Xianhong Yin, Meng Liang, Xiaoyu Liu, Meng Hao, Yi Wang

## Abstract

**Importance:** To predict the diagnosed COVID-19 patients and the trend of the epidemic in China. It may give the public some scientific information to ease the fear of the epidemic.

**Objective:** In December 2019, pneumonia infected with the novel coronavirus burst in Wuhan, China. We aimed to use a mathematical model to predict number of diagnosed patients in future to ease anxiety on the emergent situation.

**Design:** According to all diagnosis number from WHO website and combining with the transmission mode of infectious diseases, the mathematical model was fitted to predict future trend of outbreak.

**Setting:** Our model was based on the epidemic situation in China, which could provide referential significance for disease prediction in other countries, and provide clues for prevention and intervention of relevant health authorities.

**Participants:** In this retrospective, all diagnosis number from Jan 21 to Feb 10, 2020 reported from China was included and downloaded from WHO website.

**Main Outcome(s) and Measure(s):** We develop a simple but accurate formula to predict the next day diagnosis number: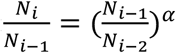,where N_i_ is the total diagnosed patient till the *i*th day, and *α* was estimated as 0.904 at Feb 10.

**Results:** Based on this model, it is predicted that the rate of disease infection will decrease exponentially. The total number of infected people is limited; thus, the disease will have limited impact. However, new diagnosis will last to March.

**Conclusions and Relevance:** Through the establishment of our model, we can better predict the trend of the epidemic in China.

## Introduction

In December of 2019, the increasing number of patients with pneumonia of unknown cause emerged in Wuhan, the capital of Hubei province, Central China. Most of the early patients had directly or indirectly related to wild animal source in the seafood and wet animal wholesale market through epidemiological survey^1-5^. Then laboratory tests found that the infection was caused by a novel coronavirus (now named as COVID-19 ^6-7^).

The spread of COVID-19 is expeditious. According to WHO statistics, the total number of diagnosed people in China was 309 in Jan 21^8^, but rapidly grew to 4537 in Jan 27^9^. Considering a simple constant exponential growth rate model^10-11^, the diagnosis number will be unimaginable soon. This triggered a major concern and motivated this study^12-14^. A mathematical model is required to predict the spread trend and emergent situation.

Over the last few decades, mathematical models of disease transmission have been useful to gain insights into the transmission dynamics of infectious diseases and the potential role of intervention strategies^15^. Epidemic transmission models focused on the spread of an infectious disease through a population. But this spread can be modeled in two fundamental ways: by looking at the entire population as a single group or set of linked subgroups (population-based) or by considering everyone that makes up the population and how interactions between unique individuals facilitate disease spread (individual-based). Common models of infectious disease included meta-population models, individual-based network models, and simple SIR-type models that incorporate the effects of reactive behavior changes or inhomogeneous mixing^16^.

Considering the limited number of data points and the complexity of the real situation, a simple but robust model is expected to work better than sophisticated epidemiology models. In this paper, we propose a robust model for next day diagnosis number predict. This model had excellent performance on prediction of infected patients in past few days. The prediction is expected to provide practical significance on social and economic application.

## Results

### Estimation of α

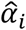 and its median aggregates were calculated (Fig.1). It was observed that 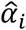 manifests a random fluctuation around a constant center. It is reasonable to assume that *α* is a constant. The estimation of *α* currently (Feb 10) is 0.904. A future diagnosis number formula is obtained based on this estimation:

**Fig. 1.**
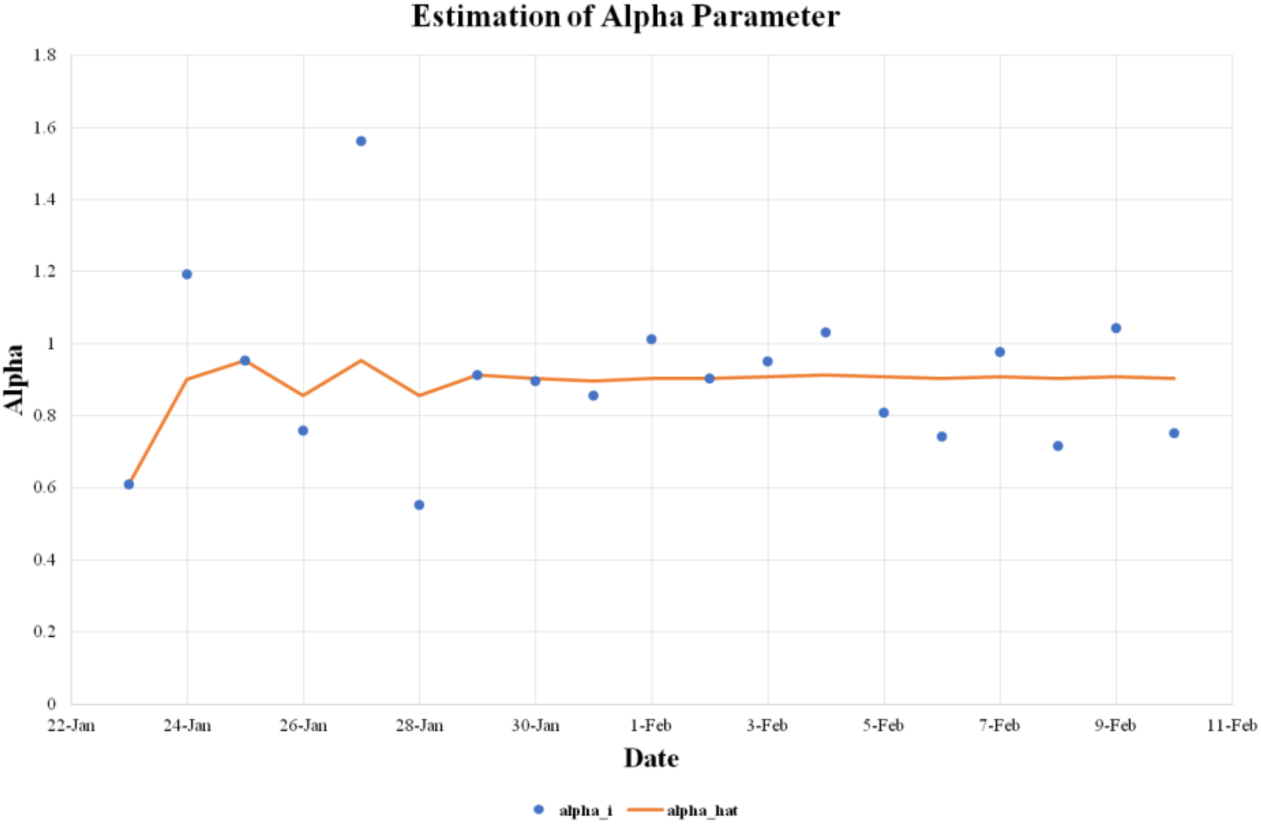
Estimation of Alpha Parameter.

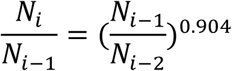

### Next Day Prediction Table

Previous numbers of diagnoses in China are used to predict the number for the next day. The prediction is started from Jan 29 as several data points are needed to build the model (Table 1). We obtain nice concordance (<2.5% error) with the real numbers next day.

**Table 1.** Observed and Predicted number of diagnosis in China

| Date | WHO | Predict | error% |
| --- | --- | --- | --- |
| 21-Jan | 309 |  |  |
| 22-Jan | 571 |  |  |
| 23-Jan | 830 |  |  |
| 24-Jan | 1297 |  |  |
| 25-Jan | 1985 |  |  |
| 26-Jan | 2741 |  |  |
| 27-Jan | 4537 |  |  |
| 28-Jan | 5997 |  |  |
| 29-Jan | 7736 | 7602 | -1.73% |
| 30-Jan | 9720 | 9760 | 0.41% |
| 31-Jan | 11821 | 11950 | 1.09% |
| 1-Feb | 14411 | 14088 | -2.24% |
| 2-Feb | 17238 | 17240 | 0.01% |
| 3-Feb | 20438 | 20269 | -0.83% |
| 4-Feb | 24363 | 23857 | -2.08% |
| 5-Feb | 28086 | 28600 | 1.83% |
| 6-Feb | 31211 | 31959 | 2.40% |
| 7-Feb | 34598 | 34335 | -0.76% |
| 8-Feb | 37251 | 37992 | 1.99% |
| 9-Feb | 40235 | 39824 | -1.02% |
| 10-Feb | 42708# | 43152 | 1.04% |
| 11-Feb |  | 45075 |  |

### New Diagnosis Number Prediction

We predict the new diagnosis number in China according to our model as Fig.2. The model predicts that new diagnosis will last to March.

**Fig. 2.**
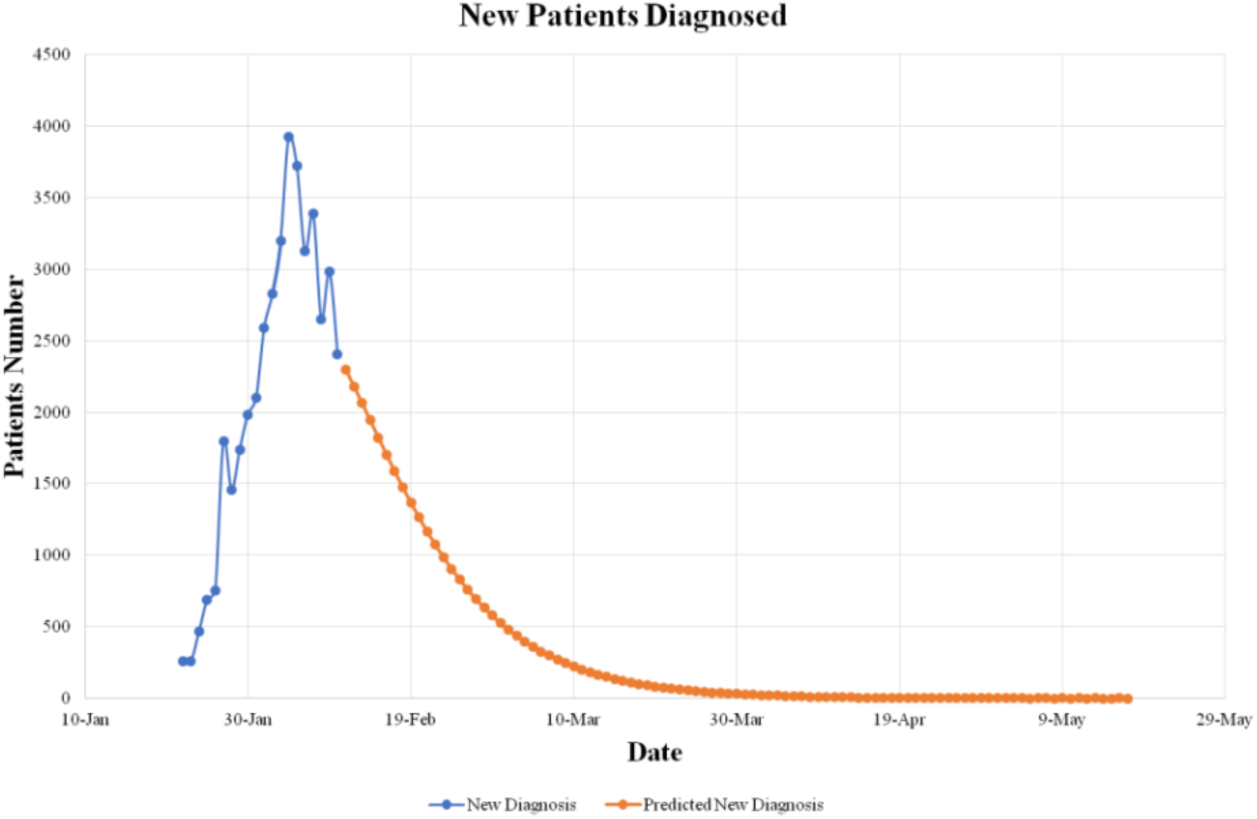
Observed (Blue) and Predicted (Orange) number of daily new diagnosis. Conclusion.

## Conclusion

Our model indicates that the COVID-19 disease infection rate decreases exponentially. This model converges to a maximum number as time increases, indicating a limited impact of COVID-19. The model also predict that the new diagnosis number will last to March, indicating that the COVID-19 disease will be well controlled.

## Discussion

We only analyzed the total number of diagnoses in China. The province scale data is available but is not as complete as total number. Here, we did not show these details analyses.

Our model is attractive as it not only predicts data accurately but also is minimal. The minimal number of parameters and robust statistics make our model robust in the real-world scenario. We develop a formula to predict the next day diagnosis number: 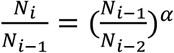,and estimate that *α* is 0.904(*α*<1) which shows that the diagnosis number of COVID-19 patients will decrease. It is very effective in controlling the spread of infectious diseases that the Chinese health authorities have taken appropriate health care actions. If the outbreak is no longer controlled, the value of *α* will be around 1 indicating that the outbreak will wreak havoc.

We obtain this elegant formula with both curve plot visualization and mathematical intuition. We plot ln (N_i_/N_i-1_) against time (Fig.3). First few observations showed a linear decay of spread rate. However, later data points realized us at Feb 4 that it is an exponential decay. Later data points further confirm our model.

**Fig. 3.**
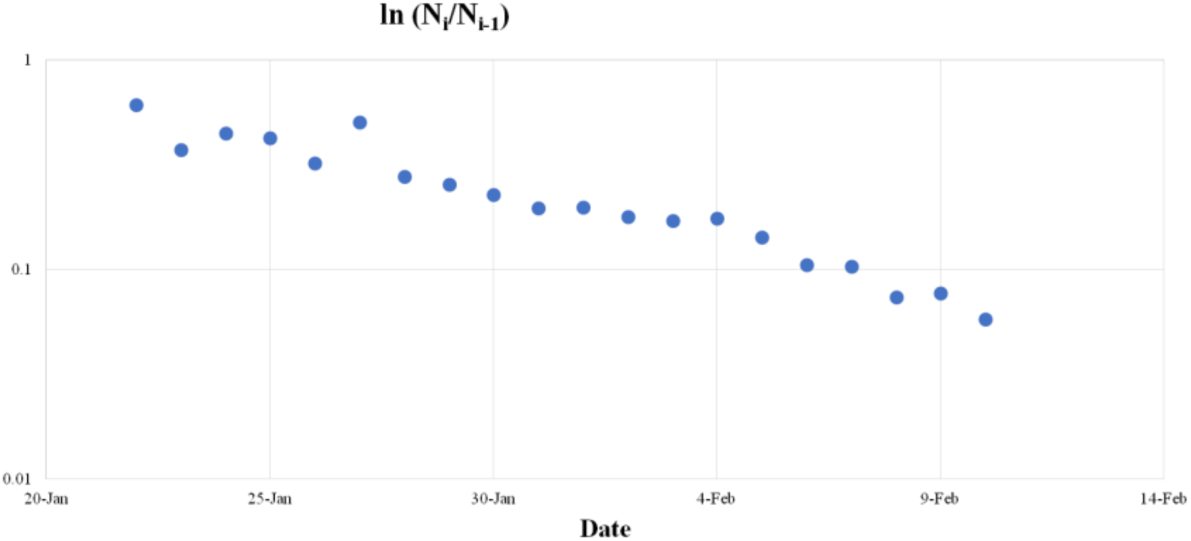
ln (N_i_/N_i-1_) against time. Reference.

## Data Availability

In this retrospective, all diagnosis number from Jan 21 to Feb 10, 2020 reported from China was included and downloaded from WHO website.

https://www.who.int/emergencies/diseases/novel-coronavirus-2019/situation-reports

## Methods

### Data Download

Daily diagnosis number of China is download from WHO situation reports (https://www.who.int/emergencies/diseases/novel-coronavirus-2019/situation-reports). The data starts from Jan 21 and currently it ends at Feb 10.

### Model

The following model is proposed based on our observations:

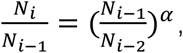

where N_i_ is the number of diagnosed patients in China according to WHO at the *i*th day, *α* is a parameter modeling the decay of diagnosis growth rate.

A simple estimation of *α* at the *i*th day would be

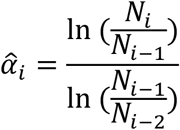

As we have multiple days of observations, historical 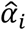 with the median statistic, which is well known for its robustness, can be aggregated.

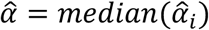

The source code of model estimation is at: https://github.com/wangyi-fudan/ncp_model

## Author Contribution

YW conceived the idea and wrote the source code. YL, XY and YW contributed data analysis, generating tables and figures, and manuscript writing. YL, XY, ML, MH, LX and YW contributed the theoretical analysis and manuscript revision. All authors contributed to final revision of the manuscript.

## Acknowledgements

We thank the Fudan University High-End Computing Center for supporting computations involved in this study.

## Reference

1. Chaolin H. et al. Clinical features of patients infected with 2019 novelcoronavirus in Wuhan, China. The Lancet. ISSN0140-6736,(2020).

2. PaulesC I. et al. Coronavirus Infections—More Than Just the Common Cold. JAMA.(2020)

3. Nishiura H. et al. The extent of transmission of novel coronavirus in Wuhan, China, 2020. Journal Of Clinical Medicine, (2020).

4. Tao L. et al. Transmission dynamics of 2019 novel coronavirus (2019-nCoV). bioRxiv 2020.01.25.919787;(2020).

5. Zhu N.et al. A Novel Coronavirus from Patients with Pneumonia in China, 2019. New England Journal of Medicine, (2020).

6. https://www.who.int/dg/speeches/detail/who-director-general-s-remarks-at-the-media-briefing-on-2019-ncov-on-11-february-2020.

7. Chua M. et al. From the frontlines of COVID-19 - How prepared are we as obstetricians: a commentary. BJOG-an international journal of obstetrics an d gynaecology. 2020 Mar 4. DOI: 10.1111/1471-0528.16192.

8. https://www.who.int/docs/default-source/coronaviruse/situation-reports/20200122-sitrep-2-2019-ncov.pdf?sfvrsn=4d5bcbca_2.

9. https://www.who.int/docs/default-source/coronaviruse/situation-reports/20200128-sitrep-8-ncov-cleared.pdf?sfvrsn=8b671ce5_2.

10. Zhao S. et al. Preliminary estimation of the basic reproduction number of novel coronavirus (2019-nCoV) in China, from 2019 to 2020: A data-driven analysis in the early phase of the outbreak. International Journal of Infectious Diseases, (2020).

11. Li Q. et al. Early Transmission Dynamics in Wuhan, China, of Novel Coronavirus– Infected Pneumonia. New England Journal of Medicine, (2020).

12. Mingwang S. et al. Modelling the epidemic trend of the 2019 novel coronavirus outbreak in China. bioRxiv 2020.01.23.916726;(2020)

13. Tianmu C. et al. A mathematical model for simulating the transmission of Wuhan novel Coronavirus. bioRxiv 2020.01.19.911669;(2020).

14. Shi Z, et al. Preliminary estimation of the basic reproduction number of novel coronavirus (2019-nCoV) in China, from 2019 to 2020: A data-driven analysis in the early phase of the outbreak. bioRxiv 2020.01.23.916395;(2020).

15. Kermack W. et.al. Contributions to the mathematical theory of epidemics: IV. Analysis of experimental epidemics of the virus disease mouse ectromelia. J Hyg (Lond);37:172–187,(1937)

16. Chowell G. et al. Mathematical models to characterize early epidemic growth: A Review. Physics of Life Reviews, S1571064516300641,(2016).

